# Pallidal evoked resonant neural activity as a biomarker for deep brain stimulation in dystonia

**DOI:** 10.1101/2025.08.05.25332536

**Authors:** Kara A. Johnson, Patricia B. Coutinho, Filipe P. Sarmento, Elizabeth Vo, Joshua K. Wong, Justin D. Hilliard, Kelly D. Foote, Coralie de Hemptinne

**Affiliations:** Norman Fixel Institute for Neurological Diseases, University of Florida, Gainesville, FL, USA; Department of Neurology, University of Florida, Gainesville, FL, USA; J. Crayton Pruitt Department of Biomedical Engineering, University of Florida, Gainesville, FL, USA; Department of Neurosurgery, University of Florida, Gainesville, FL, USA

**Author notes:** **Corresponding Author:** Kara A. Johnson, Ph.D., 3011 SW Williston Rd., Gainesville, FL 32608. **Financial Disclosure/COI:** None. **Funding:** Fixel-Tyler’s Hope Pilot Funding (PI de Hemptinne); K99 (PI Johnson).

**Keywords:** Dystonia, globus pallidus, deep brain stimulation, evoked potential, local field potential

## Abstract

**Background:** Deep brain stimulation (DBS) targeted to the globus pallidus (GP) can effectively alleviate dystonia symptoms. However, identifying optimal therapeutic stimulation parameters is challenging due to the manual programming process and the paucity of acute effects of DBS on dystonia symptoms.

**Objective:** This study aimed to investigate evoked resonant neural activity (ERNA) in the GP as a potential biomarker to guide DBS contact selection for chronic therapy in patients with dystonia.

**Methods:** In N=8 patients (N=9 hemispheres) undergoing GP DBS implantation for dystonia, intraoperative local field potential (LFP) recordings were acquired at resting-state (30 seconds) and during bursts of high-frequency stimulation delivered from each DBS contact. ERNA features (amplitude, frequency, and number of peaks) were measured and correlated with dystonia symptom severity, resting-state LFP spectral power, and postoperative chronic (12-month) therapeutic stimulation parameters.

**Results:** ERNA was consistently elicited by GP DBS but varied in amplitude, frequency, and number of peaks across individuals and stimulating contacts. Higher ERNA amplitudes were associated with stimulation at the GP internus/externus (GPi/GPe) border. ERNA frequency was negatively correlated with cervical dystonia severity (p<0.001) and resting-state alpha (8-12 Hz) power (p<0.05). In 8/9 (88.9%) hemispheres, the DBS contact that elicited the maximum ERNA matched the contact empirically selected for chronic therapy by expert clinicians through routine clinical programming.

**Conclusions:** Based on its correlation with dystonia symptom severity and therapeutic contact for chronic DBS, ERNA shows promise as an objective biomarker to improve the efficiency and efficacy of DBS for dystonia.

## INTRODUCTION

Dystonia is a complex and heterogeneous movement disorder characterized by involuntary co-contractions of muscles that cause abnormal postures or repetitive movements.^1^ Deep brain stimulation (DBS) targeted to the sensorimotor region of the globus pallidus (GP), specifically the GP internus (GPi), can be an effective therapy for reducing dystonia symptoms and improving patients’ quality of life.^2–6^ Successful DBS therapy requires postoperative programming, during which the clinician must identify stimulation parameters that effectively alleviate symptoms. However, determining the optimal parameters for dystonia DBS is complex and time-consuming because often there are minimal or no acute effects on dystonic symptoms (in stark contrast to the rapid suppression of tremor and bradykinesia in other movement disorders). It is impossible for clinicians to fully explore the thousands of possible parameter combinations, and some patients may never achieve optimal outcomes. Data-driven methods to predict the optimal stimulation parameters for DBS in dystonia are urgently needed to reduce the burden on patients and clinicians and improve the efficacy of the therapy.

One promising strategy is to use brain signals to guide DBS programming for dystonia. Recordings of local field potentials (LFPs) from the GP and other brain regions have provided insight into the pathological neural activity associated with dystonia. In particular, several studies have revealed abnormal synchronous neural activity in the theta/alpha frequency bands (4-12 Hz) in the GP and motor cortex that may be correlated with dystonia severity.^7–9^ Despite several investigations demonstrating the pathological relevance of these signals in dystonia, it remains unclear whether low-frequency power can be used to guide therapy. Other potential markers should therefore be explored to establish data-driven methods to guide DBS for dystonia.

Evoked resonant neural activity (ERNA) has emerged as a novel marker to guide DBS in movement disorders. ERNA is a high-frequency oscillatory evoked potential that occurs shortly after the DBS pulse when stimulation is delivered in the GP or the subthalamic nucleus (STN). Research has primarily focused on ERNA in Parkinson’s disease, showing that ERNA may be localized to the site of therapeutic DBS and may predict the contact used for chronic therapy with 70-80% accuracy.^10–12^ In dystonia, case reports also indicate ERNA is elicited with GP DBS,^13,14^ and a recent study showed that ERNA also occurs in the subthalamic nucleus (STN).^15^ However, ERNA in the GP has not been fully characterized in dystonia, and whether it may be used as a biomarker to guide DBS therapy remains unknown.

This study aimed to characterize ERNA in dystonia patients and determine whether ERNA might be a valuable biomarker to guide GP DBS in dystonia. We acquired intraoperative LFP recordings to elicit and record ERNA, measure its features and localization, and assess its correlation with dystonia symptom severity, other LFP biomarkers of dystonia, and empirically derived postoperative stimulation parameters. The results suggest that ERNA is a promising biomarker that could enhance the programming efficiency and efficacy of GP DBS therapy for dystonia.

## METHODS

### Patient Cohort

Patients undergoing DBS implantation surgery targeted to the GP for the treatment of dystonia at the University of Florida Norman Fixel Institute for Neurological Diseases were included in the study. Informed consent was obtained for all patients for inclusion in our institutional database (IRB #201901807).

### Deep Brain Stimulation Surgery

A DBS lead (Medtronic, USA) was implanted in the GP according to the surgical team’s standard clinical practice.^16–18^ All patients underwent awake surgery, except for one under general anesthesia. Patients were implanted with a quadripolar DBS lead (model 3387) (N=2 patients, N=2 hemispheres) or an 8-contact directional DBS lead (model B33015) (N=6 patients, N=7 hemispheres). Each patient underwent preoperative multisequence MRI, onto which a digitized Schaltenbrand-Bailey atlas was overlaid and deformed to create patient-specific anatomical segmentations. The MRI scans were then fused to a CT scan with the Cosman-Robert-Wells stereotactic frame in place. The DBS implantation trajectory was planned based on the imaging to optimize the lead location within the GP while avoiding vasculature and ventricles. Microelectrode recordings and macrostimulation via the microelectrode sheath were used to identify any necessary trajectory adjustments. Finally, macrostimulation via the implanted DBS lead was performed to ensure that stimulation-induced side effect thresholds were consistent with optimal positioning of the DBS contacts. The implanted DBS lead was then connected to an external system for simultaneous stimulation and recording (Neuro Omega, Alpha Omega, Israel).

### Intraoperative Stimulation and Recordings

Local field potentials (LFP) from the DBS contacts were recorded at 22 kHz sampling in a monopolar configuration referenced to a corkscrew electrode on the scalp. A 60-second recording with the patient at rest and off-stimulation was first acquired to measure baseline, resting-state neural activity. As described in our previous study,^10^ bursts of high-frequency stimulation were delivered twice per second for 5-10 seconds from each contact on the DBS lead sequentially from ventral to dorsal. In patients implanted with 8-contact directional leads, stimulation was delivered from each segment separately (N=6 hemispheres) or with ring mode stimulation (N=1 hemisphere). The stimulation protocol slightly varied as part of ongoing studies (N=4 hemispheres underwent constant stimulation parameters, N=3 variable stimulation frequencies, and N=2 variable stimulation amplitudes). To enable direct comparison, all analyses in the present study only included data acquired during consistent stimulation parameters across individuals [2.0 mA, 90 us, and 135 Hz (N=8 hemispheres) or 145 Hz (N=1 hemisphere)].

### Signal Processing

Signal processing was performed offline with custom Python scripts. Methods reported in our previous study^10^ were used to process the LFP recordings acquired during burst stimulation. Briefly, the recordings were high-pass filtered and bipolar referenced to reduce stimulation artifacts. For quadripolar leads, the recordings from the two contacts directly adjacent to the stimulating contact were subtracted (e.g., C1-C3 for C2 stimulation). For 8-contact directional leads, the recordings from the adjacent segmented level were averaged and then subtracted from the adjacent cylindrical contact (e.g., average of C1a/C1b/C1c-C3 for C2 stimulation). The recordings were aligned to the last pulse of each burst, and a decaying exponential was fit to the recording 4-50 ms after the last pulse and subtracted to remove amplifier baseline trends. The detrended evoked responses were then averaged across bursts. A peak-finding algorithm was applied to identify peaks and troughs to quantify the ERNA amplitude (peak-to-trough amplitude of the first peak), ERNA frequency (inverse of the time difference between the first two peaks), and the number of peaks detected. For 8-contact directional leads, ERNA features were averaged across stimulating segments for comparison with ring stimulation.

The resting-state, off-stimulation recordings were processed by selecting the first 30-second period free of noise/artifacts for subsequent analyses. The selected period was then bipolar referenced following the same methods described above and filtered using a 5^th^-order Butterworth band-pass filter (1-500 Hz). The power spectral density (PSD) from 1-500 Hz with 0.5 Hz resolution was computed using Welch’s method (SciPy welch: 1-second Hann window, 0.5-second overlap). A peak-finding algorithm (SciPy findpeaks: peaks occurring at ≥40 Hz, prominence=0.5) was implemented to identify any residual noise peaks in the PSD. Noise peaks that were detected in all contacts were then notch filtered. All peaks marked for removal were verified visually and occurred at ≥60 Hz. To account for variability in PSD offset and slope, Fitting Oscillations & One Over F (FOOOF)^19^ was used to fit an exponential from 1-100 Hz, which was then subtracted from the original PSD to obtain a normalized PSD. The normalized PSD for each contact was then averaged over canonical frequency bands: delta (1-4 Hz), theta (4-8 Hz), alpha (8-12 Hz), low beta (13-20 Hz), high beta (20-30 Hz), low gamma (30-50 Hz), and high gamma (50-100 Hz).

Each patient’s DBS electrodes were localized using previous methods^10^ and visualized relative to anatomical segmentations from the DISTAL atlas.^20^ One patient was excluded from localization due to missing postoperative CT imaging.

### Postoperative Stimulation and Clinical Assessment

Following surgery, all patients underwent standard clinical DBS programming by expert movement disorders neurologists. Stimulation parameters established at the 12-month post-surgery visit were obtained through retrospective chart review to determine whether ERNA features were correlated with the contact empirically selected for chronic therapy. Dystonia symptoms were assessed using the Unified Dystonia Rating Scale (UDRS), the Burke-Fahn-Marsden Dystonia Rating Scale (BFMDRS), and/or the Toronto Western Spasmodic Torticollis Rating Scale (TWSTRS). TWSTRS evaluations were only performed in patients diagnosed with cervical dystonia. The scores obtained at the preoperative baseline assessment and the 12-month visit were compared to assess clinical improvement. In one patient (DYS005), data were obtained from the 6-month visit due to missing 12-month data.

### Statistical Analysis

Statistical analyses were performed using Python using SciPy (version 1.10.1). Mann-Whitney-U or Wilcoxon signed-rank tests were used to compare unpaired or paired distributions. Spearman’s rank correlations were used to assess the linear correlations. Corrections for multiple comparisons using false discovery rate (FDR) were applied when evaluating correlations between ERNA features, clinical rating scale scores, and resting-state spectral power. The threshold for statistical significance was p<0.05 for all analyses.

### Data Sharing

The data that support the findings of this study are available on request from the corresponding author. The data are not publicly available due to privacy or ethical restrictions.

## RESULTS

### Cohort Characteristics

The study included a total of N=8 patients (N=6 female, mean (SD) age 55.59 (16.81) years), corresponding to N=9 hemispheres with intraoperative recordings (5 left GP, 4 right GP) (**Table 1**). The cohort comprised N=6 patients diagnosed with cervical dystonia (including N=2 with Meige syndrome), one patient with focal dystonia (right hand), and one patient with generalized dystonia (DYT-28; the only patient with positive genetic testing). The mean (SD) preoperative baseline UDRS, BFMDRS, and TWSTRS scores were 15.00 (16.81) (N=8), 13.75 (15.79) (N=8), and 45.00 (15.18) (N=5), respectively.

**Table 1.** Cohort characteristics.

| Patient ID | Sex | Type | Recording Site(s) | Anesthesia | Dystonia Medications | Baseline UDRS | Follow-up UDRS | Baseline BFMDRS | Follow-up BFMDRS | Baseline TWSTRS | Follow-up TWSTRS | Stimulation Parameters |
| --- | --- | --- | --- | --- | --- | --- | --- | --- | --- | --- | --- | --- |
| DYS001 | F | Tardive Cervical | R GPi | Awake | Tetrabenazine<br>Clonazepam | 14 | 11 | 8 | 5 | 65 | 20 | 1-C+ / 3.4 mA / 90 μs / 135 Hz |
| DYS002 | F | Cervical (Meige) | R GPi | Awake | Baclofen | 6 | 4 | 8 | 3 | - | - | 2-C+ / 2.2 mA / 90 μs / 135 Hz |
| DYS003 | M | Cervical | L GPi | Awake | - | 5 | - | 6 | 1 | 40 | 11 | 2-C+ / 2.0 mA / 60 μs / 165 Hz |
| DYS004 | M | Generalized (DYT-28) | L GPi | General | - | 55 | 21 | 52 | 10 | - | - | 1-C+ / 2.8 mA / 180 μs / 60 Hz |
| DYS005 | F | Focal | L GPi | Awake | - | 10 | 10 | 16 | 16 | - | - | 2-C+ / 1.0 mA / 90 μs / 125 Hz* |
| DYS006 | F | Cervical | L GPi | Awake | - | 6 | 5 | 6 | 6 | 39 | - | 1-2-C+ / 2.7 mA / 90 μs / 80 Hz |
| DYS007 | F | Cervical | R, L GPi | Awake | Cyclobenzaprine | 6 | 9 | 7 | 9 | 55 | - | Bilateral Interleaving<br>L1: 0-C+ / 2.6 mA / 120 μs / 125 Hz<br>L2: 1-C+ / 2.3 mA / 150 μs / 85 Hz<br>R1: 1-C+ / 1.9 mA / 120 μs / 125 Hz<br>R2: 2-C+ / 2.8 mA / 150 μs / 85 Hz |
| DYS008 | F | Cervical (Meige) | R GPi | Awake | - | 18 | 10 | 7 | 5 | 26 | 7 | 1-2+ / 2.8 mA / 120 μs / 125 Hz |
| **Total** | 6 F / 3 M | 6 Cervical / 1 Generalized / 1 Focal | 5 L GPi / 4 R GPi | 1 General / 8 Awake | 4 On Medication(s) | 15.00 (16.81) | 10.0 (5.34) | 13.75 (15.79) | 6.88 (4.70) | 45.00 (15.18) | 12.67 (6.66) |  |

### ERNA Features and Localization

Bursts of high-frequency DBS (**Fig. 1A**) elicited the characteristic ERNA response in 9/9 hemispheres (100%). The ERNA amplitude, frequency, and number of peaks were obtained from the contact with the maximum ERNA amplitude for each hemisphere. ERNA features and overall morphology varied substantially across individuals (**Fig. 1B**). The mean (SD, range) ERNA amplitude was 100.27 (58.42, 45.29-210.05) μV, ERNA frequency was 264.65 (76.40, 203.70-415.09) Hz, and number of peaks was 3.22 (1.20, 2.0-5.0) (**Fig. 1C**). ERNA features did not differ depending on the specific stimulation protocol delivered [constant (N=4 hemispheres) versus varying (N=5 hemispheres) stimulation parameters; ERNA amplitude: Mann-Whitney-U U=14.0, p=0.41; frequency: U=6.0, p=0.41; number of peaks U=6.0, p=0.37] or depending on the electrode model [quadripolar (N=2 hemispheres) versus 8-contact directional (N=7 hemispheres); ERNA amplitude: U=8.0, p=0.89; frequency: U=1.0, p=0.11; number of peaks U=6.0, p=0.88].

**Fig. 1.**
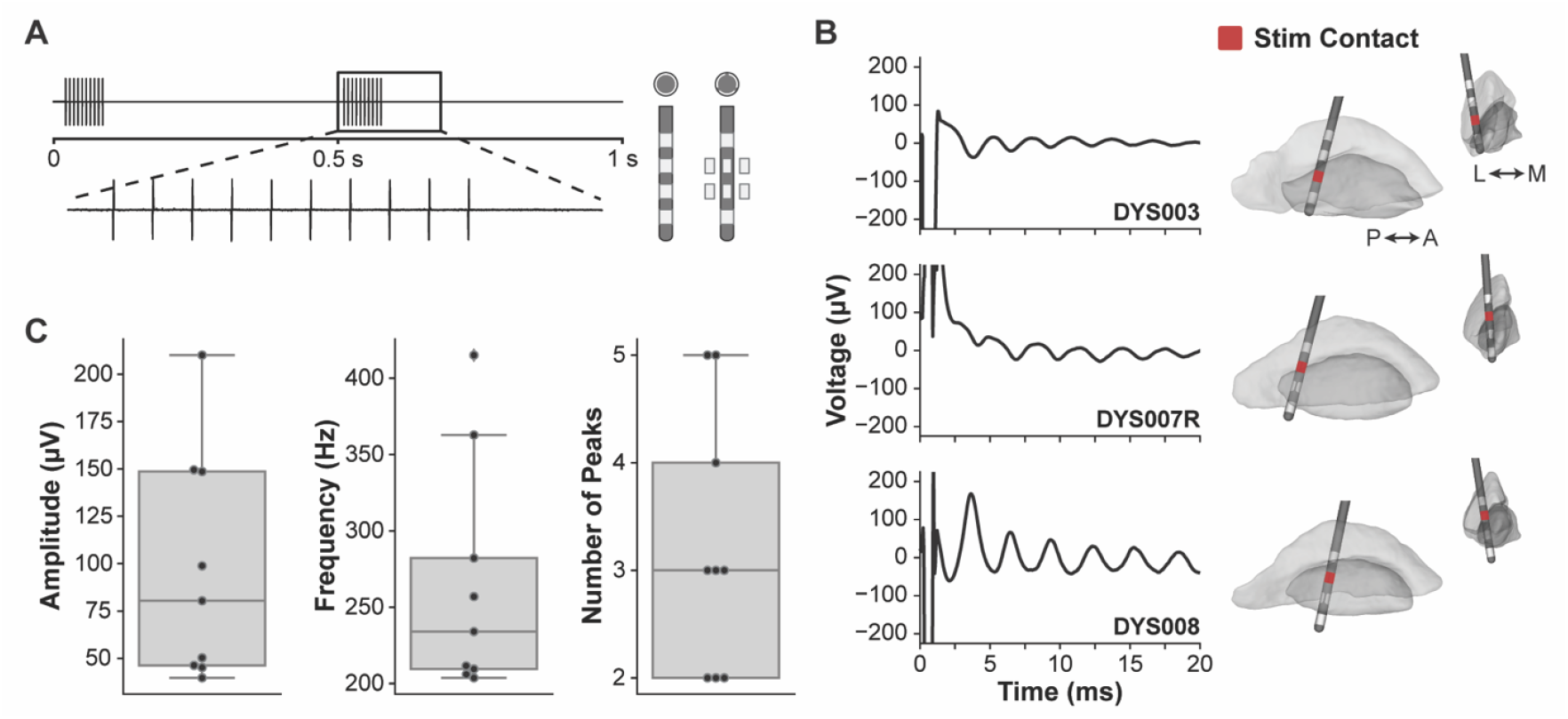
ERNA features in dystonia. **(A)** Bursts of high-frequency stimulation were delivered from each contact on the DBS electrode sequentially while recording LFP to elicit ERNA. **(B)** Representative examples of ERNA and corresponding DBS lead locations. **(C)** Distribution of ERNA features: amplitude, frequency, and number of peaks.

ERNA was observed in all hemispheres, but the features varied within individuals depending on the contact used for stimulation (**Fig. 2A-B**)—an essential characteristic if ERNA could be used as a marker of therapeutic stimulation. Only N=2 patients (N=2 hemispheres) exhibited ERNA when stimulating from only one contact and not the other. Stimulating C2 elicited significantly higher ERNA amplitude than C1 at a group level (Wilcoxon signed-rank W=1.0, p=0.031), but there was no significant difference in frequency (W=8.0, p=0.38) or number of peaks (W=6.0, p=0.68). Additionally, stimulating contacts located at the GPi/GPe border and within the GPe were associated with higher amplitude ERNA responses (**Fig. 2C**). However, there was substantial variability across patients, and precise spatial analysis was challenging in a small cohort.

**Fig. 2.**
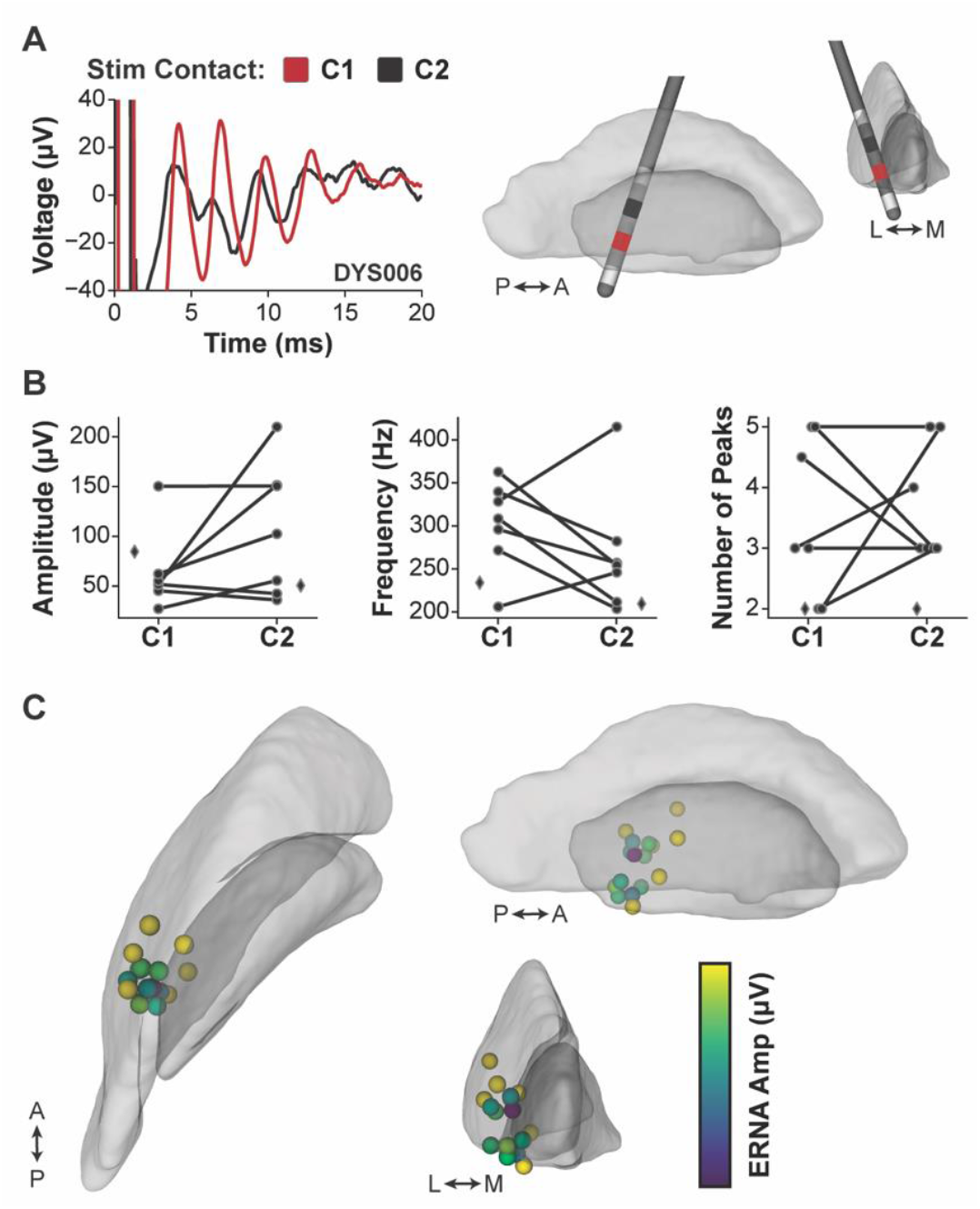
ERNA varied depending on stimulating contact and anatomical location. **(A)** Example of contrasting ERNA across stimulating contacts and the corresponding DBS electrode localization. **(B)** Distribution of ERNA amplitude, frequency, and number of peaks across contacts within individual patients. Diamonds denote individuals in which ERNA was observed with only one stimulating contact. **(C)** Stimulating contact locations across N=8 hemispheres (N=7 patients; N=16 contacts) represented by spheres and colored according to ERNA amplitude.

### ERNA Features Associated with Dystonia Severity and Resting-State Spectral Power

We investigated whether ERNA features were associated with dystonia symptom severity. ERNA frequency (from the maximum ERNA amplitude contact in each hemisphere) was negatively correlated with preoperative cervical dystonia severity (TWSTRS total score) (N=5 patients/N=6 hemispheres with available scores) (Spearman’s ρ=-0.99, p<0.001, p_FDR_=0.0027), indicating that lower ERNA frequency was associated with worse cervical dystonia symptoms (**Fig. 3A**). Notably, the one patient (DYS007) in whom bilateral recordings were acquired exhibited strikingly similar ERNA frequencies across hemispheres (left: 206.03 Hz, right: 211.54 Hz), suggesting that ERNA frequency may be an individualized marker of overall cervical dystonia severity. No other ERNA features were significantly correlated with UDRS, BFMDRS, or TWSTRS total scores.

**Fig. 3.**
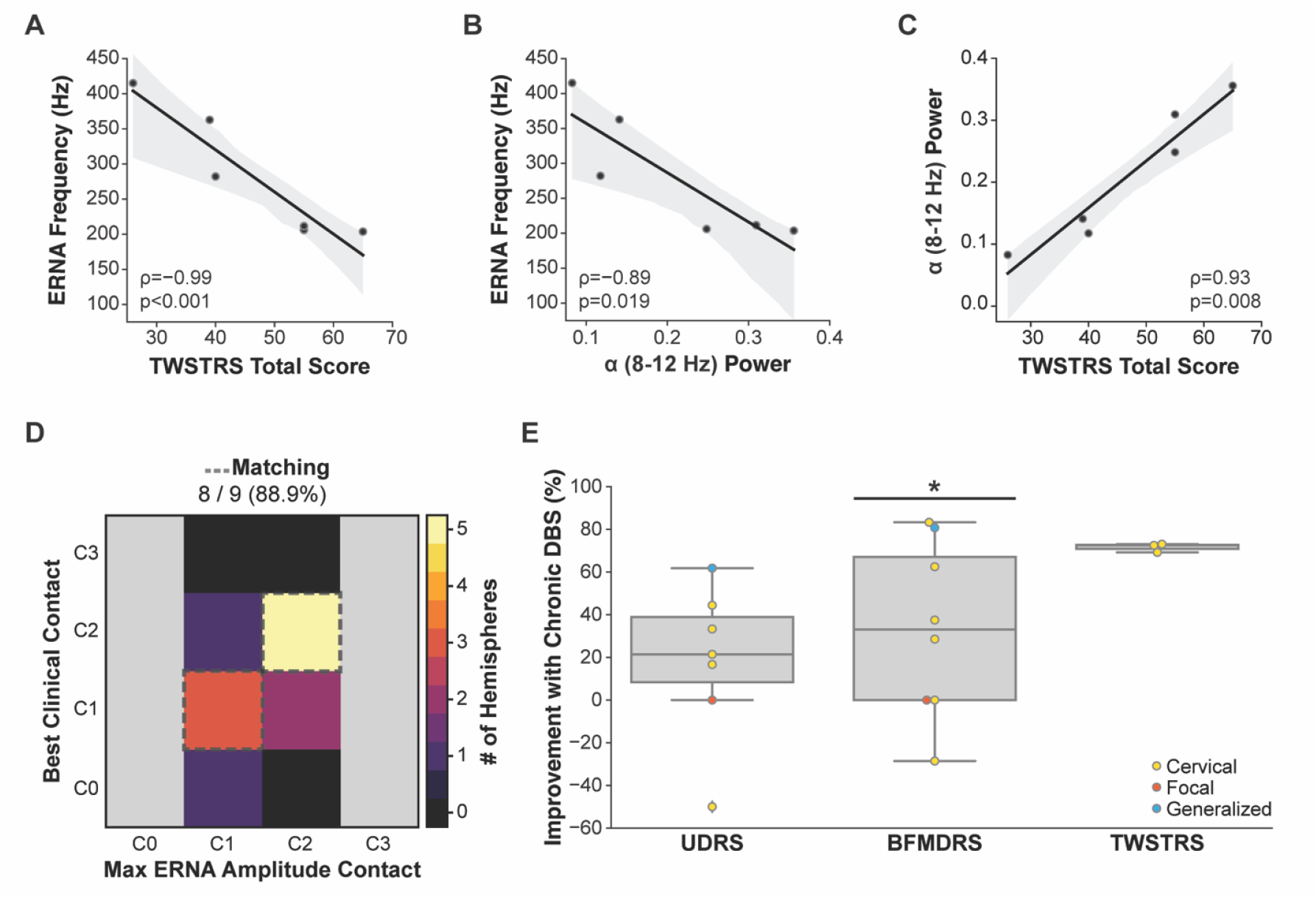
ERNA features associated with dystonia severity, resting-state spectral power, and chronic stimulation parameters. **(A)** ERNA frequency was negatively correlated with TWSTRS total score (N=5 patients / N=6 hemispheres). **(B)** ERNA frequency was negatively correlated with the normalized alpha (8-12 Hz) power. **(C)** Normalized alpha (8-12 Hz) power was positively correlated with the TWSTRS total score. **(D)** Comparison of the number of hemispheres in which the contact that elicited the maximum ERNA amplitude matched one of the contacts selected for chronic therapeutic stimulation. **(E)** Percent improvement in UDRS, BFMDRS, and TWSTRS total scores compared to preoperative baseline (colored by dystonia type) demonstrated a reduction in symptom severity (denoted by positive % improvement) across the majority of patients.

The correlation between ERNA frequency and cervical dystonia severity prompted further analysis of whether ERNA may also be related to resting-state spectral power. Within the cervical dystonia cohort, ERNA frequency (measured from the maximum ERNA amplitude contact) was indeed negatively correlated with normalized alpha (8-12 Hz) power (ρ=-0.89, p=0.019, p_FDR_=0.11) (**Fig. 3B**). We then focused on spectral power associated with symptom severity, which revealed that normalized alpha power was positively correlated with the preoperative TWSTRS total score (ρ=0.93, p=0.008, p_FDR_=0.038) (**Fig. 3C**). Normalized low gamma (30-50 Hz) power was also positively correlated with preoperative TWSTRS total score (ρ=0.81, p=0.0499, p_FDR_=0.125) but did not survive FDR correction. These findings indicate a robust relationship between lower ERNA frequency, increased alpha power, and worse preoperative cervical dystonia symptoms.

### ERNA and Chronic Therapeutic DBS

ERNA features were compared to stimulation parameters selected for chronic DBS therapy through standard clinical programming. The contact that elicited the maximum ERNA amplitude was chosen for chronic DBS therapy after parameter optimization in 8/9 (88.9%) of hemispheres (**Fig. 3D**). The only patient whose maximum ERNA contact and chronic DBS contact did not match was DYS001, whose complex pathophysiology involved tardive cervical dystonia. Chronic DBS was effective at reducing dystonia symptoms, with a mean (SD, range) improvement of 18.24% (36.09, -50.0-61.82) in UDRS scores (N=7 patients; Wilcoxon signed-rank comparing percent improvement distribution to 0 (no improvement), W=16.0, p=0.12), 33.01% (40.87, -28.57-83.33) in BFMDRS scores (N=8 patients; W=19.5, p=0.047), and 71.60% (2.07, 69.23-73.08) in TWSTRS scores (N=3 patients; W=6.0, p=0.13) (**Fig. 3E**). The effect of DBS on UDRS and TWSTRS did not reach statistical significance, likely because the sample size is small, and the UDRS/BFMDRS do not purposely capture cervical dystonia symptoms. However, detailed clinician notes reported beneficial outcomes for all patients except for two; DYS005 unfortunately continued to experience severe focal hand dystonia with insufficient response to DBS, and there was some uncertainty about a neurodegenerative condition. The second was DYS007, whose head tremor and pain were improved but not head posture. Interestingly, a higher ERNA frequency corresponded to greater improvement in symptoms; however, this exploratory analysis only included N=3 patients.

## DISCUSSION

This study investigated ERNA in the GP as a potential biomarker to guide DBS therapy in patients with dystonia using intraoperative neural recordings and retrospective analysis of clinical data. The results revealed that ERNA: (1) was consistently recorded from the GP in patients with dystonia; (2) varied substantially across individuals and stimulating contacts; (3) was associated with preoperative dystonia symptom severity and resting-state alpha power; and (4) predicted the DBS contact selected for chronic therapeutic stimulation. Altogether, our findings provide compelling evidence that ERNA in the GP could serve as an objective biomarker to improve the programming efficiency and overall efficacy of DBS for dystonia.

### ERNA as a Marker of Basal Ganglia Network Engagement

Most of our understanding of ERNA has come from studies performed in patients with Parkinson’s disease undergoing DBS targeted to either the STN^11,21–23^ or the GP^10,24^. These investigations defined ERNA characteristics and found that ERNA may be localized to the DBS contact and target location most effective for alleviating Parkinson’s disease motor symptoms.^10,12,23^ The current hypothesis is that ERNA is generated through modulation of reciprocal excitatory and inhibitory connections between the STN and the GP (particularly the GPe),^25^ which is supported by computational models of basal ganglia network dynamics^21^ and simultaneous stimulation/recording from both the GP and STN.^13^

A few studies indicate that ERNA also occurs in patients with dystonia, suggesting ERNA is not specific to Parkinson’s disease but rather a biomarker of basal ganglia network engagement. The studies thus far have provided a preliminary basis for understanding ERNA in dystonia, including one study in the STN,^15^ case studies in the GP^14^ and simultaneous GP-STN DBS,^13^ and a recent study on ERNA using paired-pulse DBS in the GP or STN (including one patient with dystonia).^24^ To our knowledge, we present the first study using high-frequency DBS to characterize GP ERNA in a cohort of patients with dystonia and determine its potential clinical utility. Our results suggest that ERNA in dystonia exhibits similar characteristics to ERNA in Parkinson’s disease. However, direct comparisons across the two disorders will need to be performed.

Beyond ERNA, other studies have investigated evoked potentials to uncover the multiscale mechanisms of DBS in dystonia and guide treatment. At the local synaptic scale, GP DBS elicited field evoked potentials that may reflect pallido-striatal activity, which exhibited both short- and long-term plasticity during and after high-frequency DBS, respectively, and were reduced in dystonia compared to Parkinson’s disease.^26,27^ At the subcortical-cortical network scale, GP DBS has been shown to elicit evoked potentials in the primary motor cortex, thought to reflect pallido-thalamo-cortical activity, which may correlate with stimulation in the GPi and the effective contact for therapeutic DBS.^28–30^

Altogether, both local evoked potentials, such as ERNA, and cortical evoked potentials provide complementary information about the network mechanisms of DBS. Further research comparing and contrasting these signals is needed to understand their relationship and relative predictive power to guide DBS therapy in dystonia.

### Relationship Between ERNA Frequency, Dystonia Symptoms, and Alpha Power

Beyond a biomarker of basal ganglia network engagement, our data also indicate that ERNA features may reflect neurophysiological underpinnings of cervical dystonia. We found a strong correlation between ERNA frequency, alpha (8-12 Hz) spectral power, and cervical dystonia symptoms, where lower ERNA frequency and increased alpha power correlated to worse symptoms. In line with our results, several previous studies have implicated low-frequency power (theta/alpha, 4-12 Hz) in the GP in dystonia pathophysiology (summarized in a recent review^31^). Low-frequency power may be suppressed by therapeutic DBS^32,33^ and localized to the site of effective stimulation in dystonia.^8^ In addition to local recordings in GP, low-frequency power and coupling of neural activity have also been observed in cortical^9^ and simultaneous GP-cortical recordings^7,34^, suggesting that dystonia pathophysiology involves excessive low-frequency oscillations across basal ganglia-thalamo-cortical networks.

Although our results implicating low-frequency power are supported by previous research, the mechanism underlying the correlation between ERNA frequency and alpha power remains unclear. Interestingly, in Parkinson’s disease, higher ERNA frequency has been correlated with increased beta (13-30 Hz) power, a biomarker of akinetic symptoms in PD.^35^ Our correlation was in the opposite direction, where lower ERNA frequency corresponded to increased alpha power. One hypothesis is that spontaneous oscillatory activity may mirror the dynamics of the GP-STN circuit in response to stimulation. This would mean that, in Parkinson’s disease, elevated spontaneous higher-frequency power (i.e., beta) shifts the propensity of the GP-STN circuit to respond to stimulation at a higher ERNA frequency. In contrast, in dystonia, elevated lower-frequency power (i.e., alpha) shifts the propensity of the GP-STN circuit to respond to stimulation at a lower ERNA frequency. However, understanding the causality and mechanisms of the relationship between these signals requires further experiments.

### Guiding DBS Parameter Selection and Future Technology in Dystonia

Identifying therapeutic DBS parameters in dystonia is challenging due to the heterogeneity in symptoms, lack of objective biomarkers, and symptom response to DBS can take several months to manifest fully.^36^ Identifying objective biomarkers to guide DBS programming could expedite the process and potentially improve outcomes. ERNA showed a promising correlation with the contact selected for chronic therapeutic DBS with 88.9% accuracy. Another case study using paired pulse stimulation also found that ERNA correlated with the contact used for therapeutic DBS in a patient with generalized dystonia.^14^ Future protocols could further refine this prediction by exploring the effect of varying DBS parameters on ERNA to predict not only the stimulation contact, but also the amplitude, pulse width, and frequency.

One potential clinical application of low-frequency power and ERNA is their utility as feedback signals for closed-loop DBS.^37^ Most of the studies of LFP biomarkers in dystonia have been performed in intra-, perioperative, or clinic environments. However, the advent of clinical DBS devices capable of chronic neural recordings has enabled new insight into the naturalistic fluctuations in GP activity, dystonia symptoms, and response to DBS therapy.^38–41^ Although current devices are not equipped with the sampling rate required for detecting ERNA (200-500 Hz oscillatory signal), future iterations could be used to investigate whether ERNA might be a valuable biomarker to track dystonia symptoms chronically or control closed-loop DBS algorithms.

ERNA may also be a viable biomarker to guide intraoperative targeting of GP DBS, given its localization to the GPi and GPi/GPe border.^10^ Such a biomarker could especially be impactful during MRI-guided asleep DBS surgery. Importantly, robust ERNA recordings were obtained in one patient in our cohort who underwent asleep DBS surgery. Another study has also shown ERNA is reliably detected under general anesthesia in patients with Parkinson’s disease^42^, providing evidence that ERNA may be feasible to aid in target confirmation.

## Limitations

The present study included a small cohort, although fewer patients undergo DBS for dystonia than for other movement disorders. The patients included predominantly those diagnosed with cervical dystonia, which prevented comparing ERNA across subtypes; however, we plan to investigate in future studies. Some patients were taking dystonia medications, which may affect neural activity in the GP and should be explored in a larger cohort. Our analysis was simplified to a single set of stimulation parameters, and some stimulation sites may require higher stimulation amplitudes to induce ERNA. Bipolar referencing required to mitigate stimulation artifacts limited our analysis to only the two middle contacts of the DBS lead; however, patients at our center are most commonly programmed on these contacts (shown in **Table 1**). The analyses of clinical outcomes and DBS programming data were retrospective, which may be biased due to variability in clinical assessment. Prospective studies will be necessary to validate the predictive power and clinical utility of ERNA to guide DBS in dystonia.

## Conclusions

Optimizing GP DBS therapy for dystonia is challenging due to disease heterogeneity, minimal acute symptom response to stimulation, and an unclear understanding of the most effective stimulation site. Our findings demonstrate that ERNA, which has been predominantly studied in Parkinson’s disease, is also elicited by GP DBS in dystonia. ERNA was associated with the DBS contact selected for chronic therapy with high accuracy (88.9%), and ERNA frequency may reflect cervical dystonia symptom severity in conjunction with spontaneous resting-state alpha power. Given its correlation with effective chronic DBS parameters and disease severity, ERNA shows promise as an objective biomarker to guide DBS therapy for dystonia.

## ACKNOWLEDGMENT

We gratefully acknowledge the Norman Fixel Institute for its continued support of interdisciplinary research in movement disorders, and we are thankful to the patients and families who generously contribute their time and data.

## AUTHORS’ ROLES

(1) Research Project: A. Conception, B. Organization, C. Execution, D. Supervision; (2) Data acquisition: A. Patient recruitment, B. Execution; (3) Statistical Analysis: A. Design, B. Execution, C. Review and Critique; (4) Manuscript Preparation: A. Writing of the First Draft, B. Review and Critique.

KAJ: 1A, 1B, 1C, 2B, 3A, 3B, 4A. PBC: 1C, 2B, 4B.

FPS: 1C, 2B, 4B.

LV: 1C, 4B.

JKW: 3C, 4B.

JDH: 2A, 2B, 4B.

KDF: 2A, 2B, 4B.

CDH: 1A, 1D, 3C, 4B.

